# Phylogenomics and phylodynamics of SARS-CoV-2 retrieved genomes from India

**DOI:** 10.1101/2020.06.23.20138222

**Authors:** Sameera Farah, Ashwin Atkulwar, Manas Ranjan Praharaj, Raja Khan, Ravikumar Gandham, Mumtaz Baig

## Abstract

The ongoing SARS-CoV-2 pandemic is one of the biggest outbreaks after the Spanish flu of 1918. Understanding the epidemiology of viral outbreaks is the first step towards vaccine development programs. This is the first phylodynamics study attempted on of SARS-CoV-2 genomes from India to infer its current evolution in the context of an ongoing pandemic. Out of 286 retrieved SARS-CoV-2 whole genomes from India, 138 haplotypes were generated and analyzed. Median-joining network was built to investigate the relatedness of SARS-CoV-2 haplotypes in India. The BDSIR package of BEAST2 was used to calculate the reproduction number (R0) and the infectious rate of the virus. Past and current population trend was investigated using the stamp date method in coalescent Bayesian skyline plot, implemented in BEAST2 and by exponential growth prior in BEAST 1.10.4. Median-joining network reveals two distinct ancestral clusters A and B showing genetic affinities with Wuhan outbreak sample. The network also illustrates the autochthonous development of isolates in a few instances. High basic reproduction number of SARS-nCoV-2 in India points towards the phase of active community transmission. The Bayesian skyline plot revel exponential rise in the effective population size (*N*_*e*_) of Indian isolates from the first week of January to the first week of April 2020. More genome sequencing and analyses of the virus will be required in coming days to monitor COVID19 after the upliftment of lock down in India.

## Introduction

In human history, viral outbreaks have been omnipotent threat to global human health right from the spreading of epidemic Spanish flu in 1918 to the recent coronavirus pandemic in December 2019^1,2,3^. Coronaviruses are the members of family Coronaviridae with the characteristic crown-like structure, having 26 to 32 kb of positive sense single-stranded RNA genome, while SARS-CoV-2 has a genome of 29 844 to 29 891 nt^4,5,6^. Prevalence of coronaviruses (CoVs) has been reported previously in livestock and wild species^7, 8,9,10^. Their rapidly evolving nature makes them capable to adapt and infect a range of hosts with broad tissue tropism ^11, 12^. The first corona viral outbreak - porcine epidemic diarrhea (PED), was reported in pigs in Europe and Asia in 1971^8^. However, the zoonotic shift of CoVs in humans began with the worldwide outbreak of SARS-CoV-1 in China during 2002-03 and subsequently with the Middle East respiratory syndrome (MERS) in 2012^13, 14, 15^. In the current ongoing epidemic, the first case of the novel coronavirus (nCoV), caused by the new strain of coronaviruses i.e. the SARS-CoV-2 was reported from the Wuhan city of China on 31 December 2019 and spread worldwide by January 2020^16,17^. World Health Organization declared this outbreak as a global pandemic on 11^th^ March 2020. The novel coronavirus strain is characterized primarily by its potential to infect the respiratory tract of human, causing severe pneumonia in the affected person. The whole genome sequencing and phylogenetic analysis suggests that bats might be the original hosts ^5^. Several studies, however, point towards, S gene of SARS-CoV-2 exhibiting high similarity of functional domains with isolates from the pangolin^18, 19,20^. Advances in computational genomics and the availability of genomic data enable us to understand the spatial distribution, and epidemiology of pandemics. The whole-genome sequencing and annotation of novel SARS-CoV-2 reference genome opens up rapid sequencing and assembly of many global SARS-CoV-2 genomes worldwide. Pathogen genomics is an effective tool not only in monitoring the ongoing pandemic but also in effective vaccine development. Genomic data contributed by various labs across the world provides a platform to track down the origin and events of community transmission of the virus in their countries^21^. Genomic studies already proved crucial in contact tracing of infection and would become more important in the second wave of infection after the release of lockdown in the ongoing covid19 pandemic^22^. In this study, we analyzed 121 whole genomes of SARS-CoV-2 from India to infer, (i) phylogeography of SARS-nCoV-2 within India, (ii) infection genomics to estimate reproduction number and infection rate and (iii) the past and present evolutionary trajectory of SARS-CoV-2.

## Materials and Methods

### Multiple sequence alignment and haplotype selection

Whole genome sequence information on SARS-CoV-2 isolates from India, available in GISAID database until 4^th^ May 2020, was retrieved. Out of 286 SARS-CoV-2 genomes, samples with missing information and gaps were discarded to arrive at a total of 219 Indian SARS-CoV-2 genomes. These genomes were aligned using Clustal Omega^23^ and number of haplotypes was determined using DnaSP v6^24^. Based on the emergence of 138 haplotypes, the resulting dataset was further used for downstream analyses.

### Median Joining Network

In evolutionary study, phylogenetic network becomes a method of choice for reconstructing evolutionary paths in many species. Median-joining Network (MJN) is one such algorithm developed to reconstruct the unambiguous evolutionary history of species^25^. Median-joining Network (MJN) was constructed with 138 genomes covering all major states of the India (Supplementary table 1).

### Estimation of effective reproductive number and phylodynamics

Birth-death serial model (BDSIR) as implemented in the BEAST2 package was used to estimate the effective reproduction number of SARS-CoV-2 in India^26, 27^. In BEAST2, HKY nucleotide substitution model with gamma category count 4, relaxed lognormal clock with clock rate of 8.3E-5 subs/site/month corresponding to 1×10^−3^ subs/site/year^25^ were applied. In MCMC analysis, parameters were sampled at every 1000 generations over a total of 10 million generations. Basic reproduction number (R0) and BecomeUinfectiousRate were set with the mean distribution to 18.00, assuming the meantime of recovery between 18 to 20 days. BEAST v1.10.4 with similar settings of HKY nucleotide substitution model with gamma category count 4, relaxed lognormal clock and a clock rate of 8.3E^-5^ subs/site/month corresponding to 1×10^−3^ subs/site/year^28^ was utilized to reconstruct the evolutionary dynamics of SARS-CoV-2. The tree prior was set on the coalescent exponential growth to calculate the growth rates of the virus in India. The effective sample sizes and 95% highest posterior density intervals for the parameters like basic reproduction number, BecomeUinfectiousRate, growth rate and the demographic reconstruction of growth rates with exponential growth priors were inspected using Tracer v1.7.0. The trees file was summarized in TreeAnnotator by setting the burnin percentage to 10 and the target tree type to maximum clade creditability tree, while the node heights were set to mean heights. The time-scaled Maximum Clade Credibility (MCC) tree based on MCMC analysis of the 138 SARS-CoV-2 genomes was visualized in FigTree v1.4.4 tree viewer.

### Population dynamics of SARS-CoV-2

Bayesian Skyline Plot (BSP) method as implemented in BEAST v2.2.0 was used to estimate the effective population size (*N*_*e*_) for these 138 Indian isolates. Stamped-date method with HKY nucleotide substitution as model coupled with 4 gamma category and coalescent Bayesian skyline tree priors were set for the analysis. The Markov Chain Monte Carlo (MCMC) chain length of 10 million steps was applied and first 10% were discarded as burn-in and strict clock rate of 8.33E^-5^ subs/per/site/per month was used. The log file and tree log file were analyzed to draw the BSP in Tracer v1.7.0.

## Results and discussion

The Median-Joining Network showed occurrence of ancestral clusters alongside their newly mutated daughter clusters and haplotypes. The network was defined by two main clusters marked as “A” and “B” (Fig 1). Both A and B clusters show linkage with the Wuhan, China outbreak haplotype (EPI_ISL_406798) of 26^th^ December 2019. The cluster A, which is dominant in Gujarat differed by a single median vector from the Wuhan haplotype as compared to cluster B, dominant in south India, which differed by two median vectors. In a biological network, median vectors are interpreted either as unsampled or extinct individuals. Based on this relationship, cluster A was considered as the ancestral node. Genomes of unknown origin contributed by NIV, Pune showed more affinity with SARS-CoV-2 genomes from Wuhan and Ladakh, and was linked to ancestral cluster A. However, genome isolates from West Bengal illustrated affinities with both clusters. Of note, haplotypes from worst hit Maharashtra state including Mumbai, exhibit closer relatedness to cluster B. Further, the SARS-CoV-2 haplotypes from Delhi are more widespread in distribution, while those from Madhya Pradesh display more closeness to ancestral cluster A. Many daughter haplotypes accumulated 1 to 4 mutations that were derived from cluster A and B. At one instance in the network, a daughter cluster derived from cluster B illustrated sharing of haplotypes from Delhi, Telangana and Assam and can potentially be considered as third minor sub cluster. This sub cluster further showed emergence of newer haplotypes by accumulating mutations in the range of 2-3 with sole representative from Gujarat. Based on this median joining network, autochthonous and community transmission of virus in India cannot be ruled out. Similarly, the Time-scaled Maximum Clade Credibility (MCC) tree based on MCMC analysis with an Exponential Growth tree prior reveals the basal position of haplotype from Kerala (Fig 2). The tree shows the splitting of haplotypes into two major clades i.e. Telangana and Gujarat, whereas the samples contributed by National Institute of Virology (NIV), Pune, Maharashtra displayed monophyletic status with the isolates from Telangana (Fig 2). The effective reproduction number (R0) in the SARS-CoV-2 for India was found to be 3.683 (95% HPD interval: 2.411, 5.401). The estimated mean infectious rate, which is the actual time needed for recovery after the infection, was estimated as 7.44 (95% HPD interval: 4.51, 9.99), which roughly corresponds to ∼40 to 50 days. The current population dynamics of SARS-CoV-2 in India from the last week of January (27^th^ January 2020) to the first week of May (4^th^ May 2020) was plotted using Bayesian Skyline Plot. Population dynamics of Indian isolates exhibited sigmoidal type of distribution, with exponential growth in effective population size starting from last week of January to second week of February (Fig 3A). The pandemic peaked in India around 2^nd^ week of February, plateaued around last week of February to 4^th^ May 2020. However, growth rate curve reconstruction using exponential growth tree prior depicts continuous increase in effective population from last week of January till 4^th^ May 2020 (Fig 3B).

**Fig 1.**
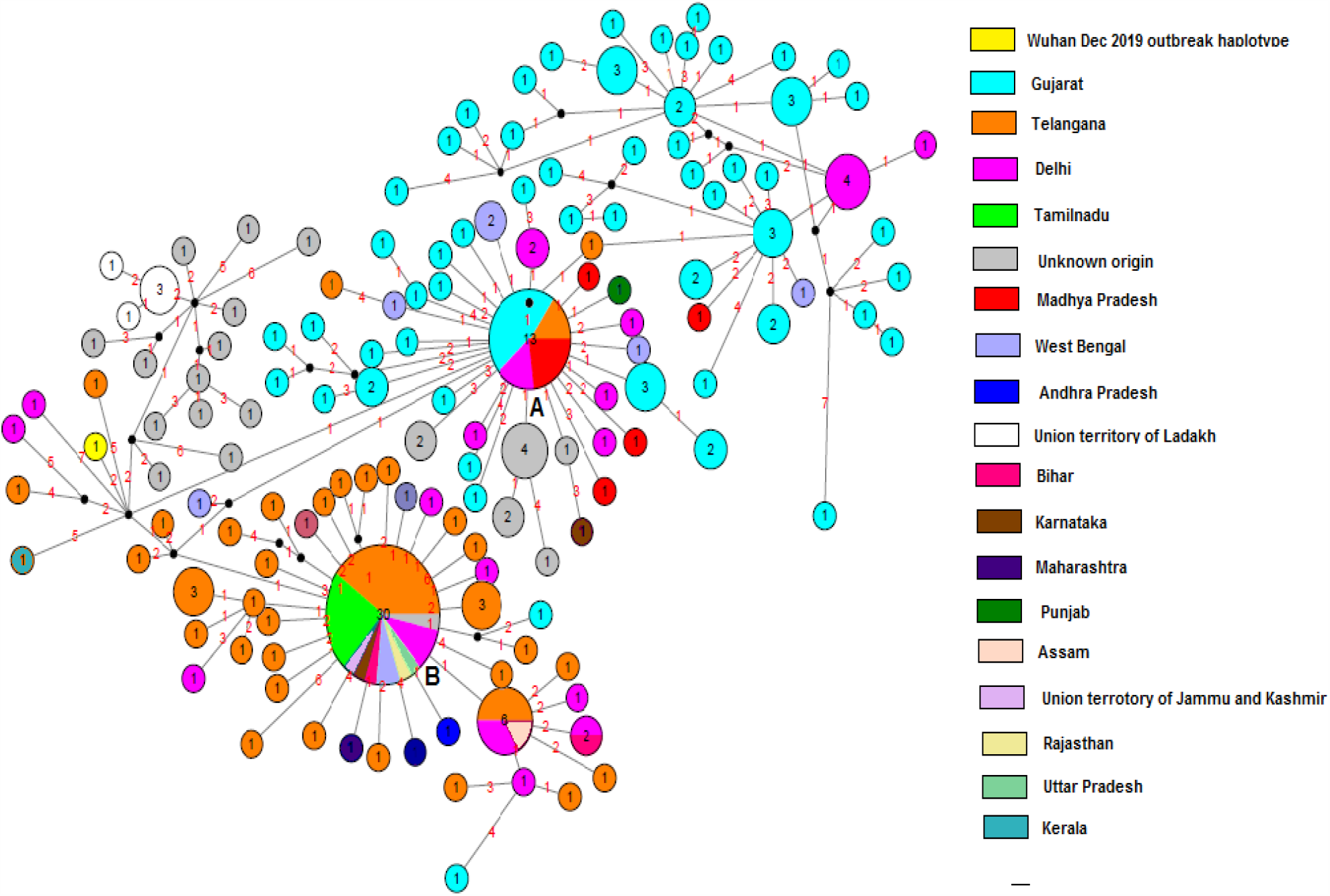
Median-Joining Network constructed out of 138 Indian haplotypes shows occurrence of ancestral clusters alongside their newly mutated daughter clusters and haplotypes. The median joining network inferred two main clusters marked as “A” and “B” showing linkage with the Wuhan, China outbreak haplotype”. Circle areas are proportional to the number of shared haplotypes, and numbers inside circle illustrate number of haplotypes, each number in red on the links represents number of mutated nucleotide position. The sequence range under consideration is 56 to 29,797, with nucleotide position (np) numbering according to the Wuhan, China EPI_ISL_406798 reference sequence. For MJN, Network5011CS (https://www.fluxusengineering.com/) was used with the parameter epsilon set to zero.

**Fig 2.**
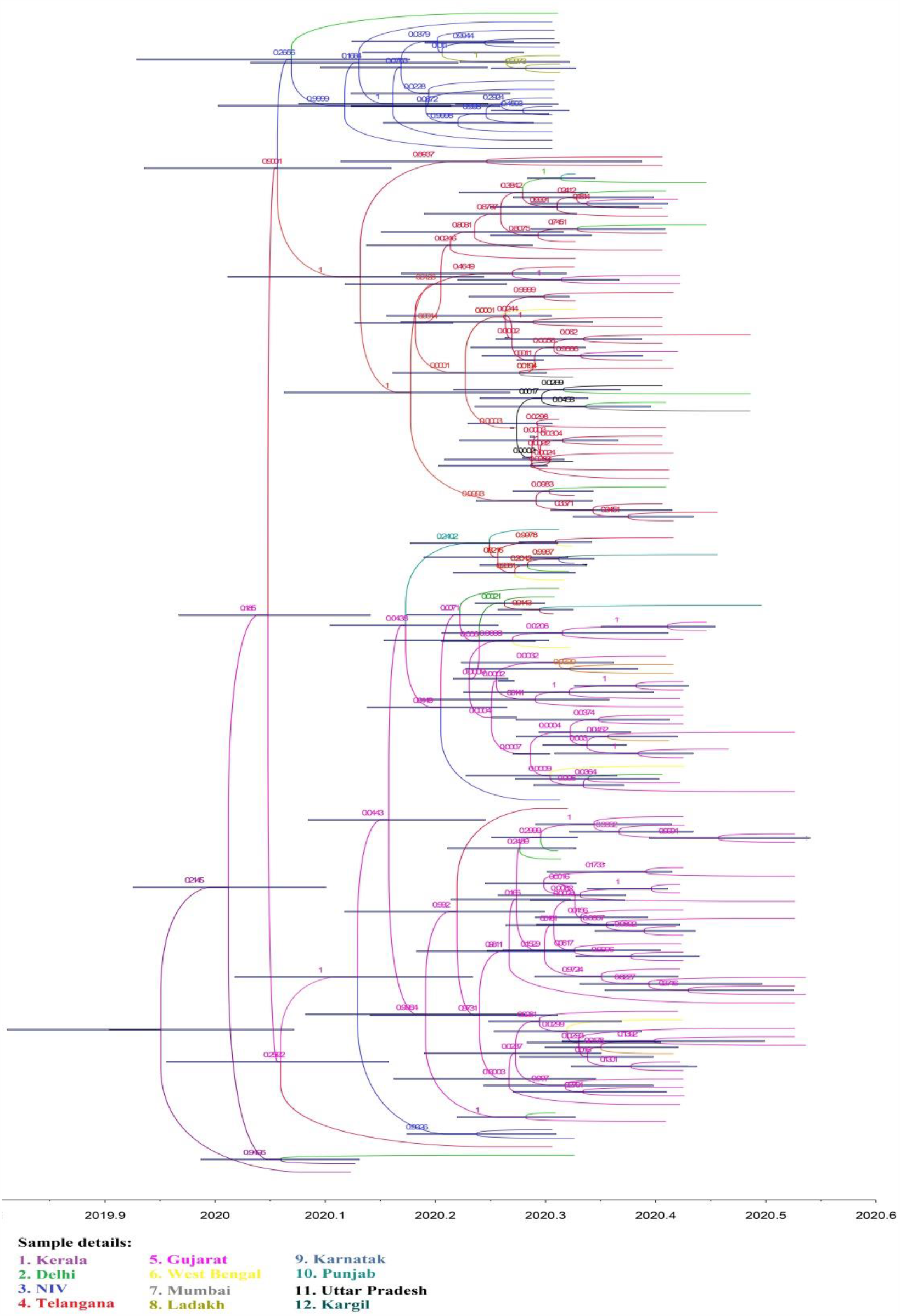
The Maximum Clade Credibility (MCC) tree based on MCMC analysis of the 138 Indian SARS-CoV-2 genomes with an Exponential Growth tree prior.

**Fig 3.**
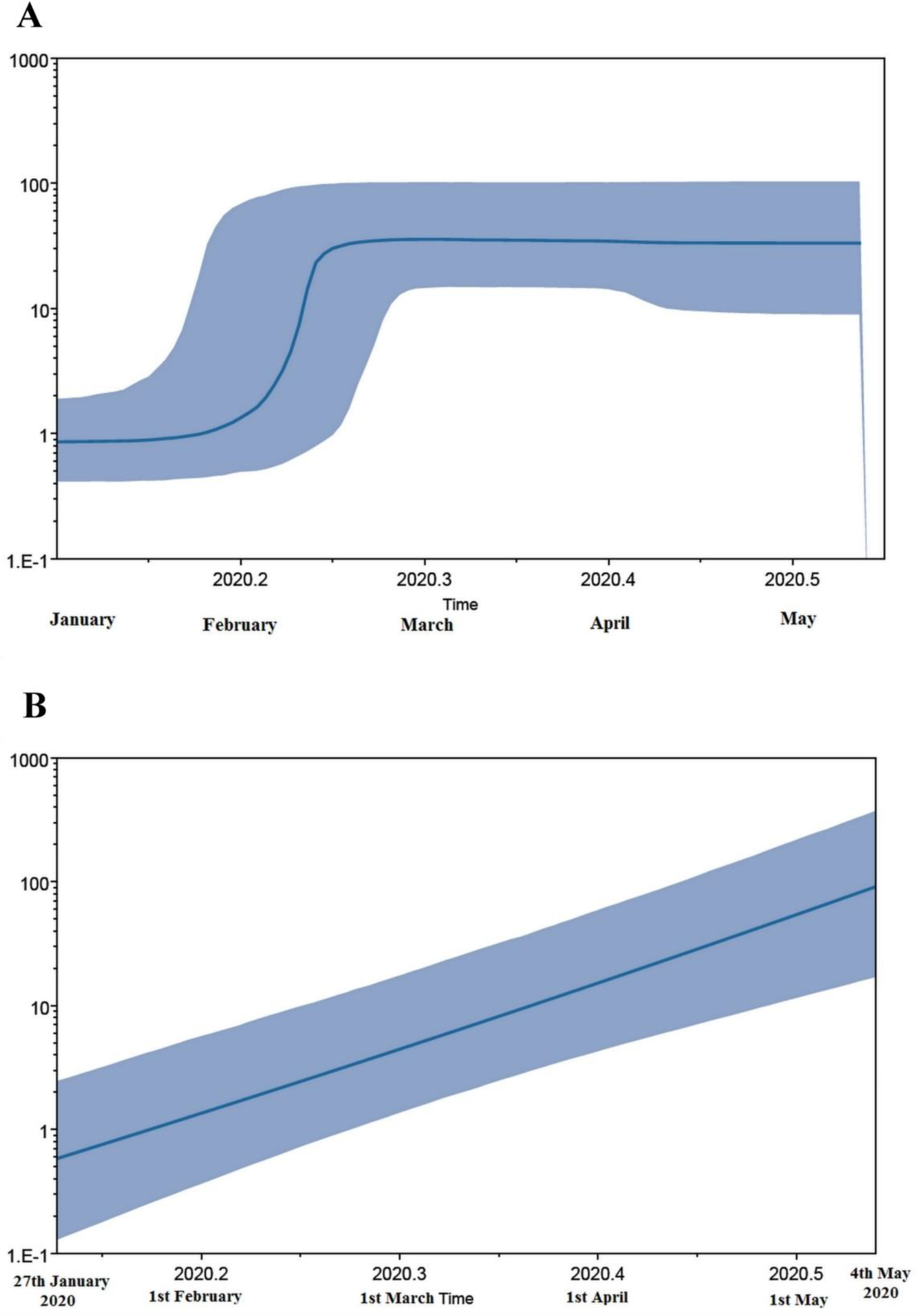
Bayesian Skyline Plots (Fig A and B) constructed using 138 Indian isolates by both coalescence Bayesian skyline and exponential growth tree priors depicts the effective population size (*Ne*) on the Y-axis, while the X-axis denotes the timeline in months.

Our phylodynamic analysis confirms the high effective reproduction number and deaths recorded in India during this period. Most probably, the occurrence of plateau phase in the Bayesian skyline plot after the last week of February resulted due to the nationwide lockdown and social distancing measures (Fig 3A). Likewise, some findings also suggest that the exposure to high temperature also contributed towards the lowering of activity and life span of SARS-CoV-2^29, 30,31^. In another recently published study, role of physical distancing measures taken in Wuhan, China beginning April played significant role in lowering the reproductive number of virus^32^. The exponential growth was also confirmed in the population dynamics, which congruent with the continuous rise of SARS-CoV-2 infections in India.

To conclude, our study provides baseline genome-based phylodynamic information, highlighting genetic affinities between the viral isolates, sequenced from the major states of the India. In coming days, sequencing and analyses of more numbers of SARS-CoV-2 genomes from India would help in dealing with the second wave of community transmission after the relaxation of lockdown. At the same time, genomic information produced through such studies can also be utilized to fill the gaps created due to unrealistic assumptions, lack of contact tracing, sampling errors and limited diagnostic testing.

## Data Availability

The dataset of the sequences used in this study is provided with the manuscript.

## Acknowledgements

We gratefully acknowledge all authors and submitting laboratories of the sequences from GISAID. Sameera Farah and Mumtaz Baig are thankful to Prof. Elizabeth Boulding and Compute Canada for providing access to high computation facilities at www.cedar.computecanada.ca

## References

1. Patterson KD, Pyle GF. The geography and mortality of the 1918 influenza pandemic. Bulletin of the History of Medicine. 1991; 65 (1): 4–21.

2. Spreeuwenberg P, Kroneman M, Paget J. Reassessing the Global Mortality Burden of the 1918 Influenza Pandemic. American Journal of Epidemiology. 2018; 187(12): 2561– 67. doi:10.1093/aje/kwy191

3. Biondi-Zoccai G, Landoni G, Carnevale R, Cavarretta E, Sciarretta S, Frati G. SARS-CoV-2 and COVID-19: facing the pandemic together as citizens and cardiovascular practitioners. Minerva Cardioangiologica 2020 Mar 9. DOI: 10.23736/S0026-4725.20.05250-0.

4. Woo PC, Huang Y, Lau SK, Yuen KY Coronavirus genomics and bioinformatics analysis. Viruses 2010; 2:1804–20.

5. Lu R, Zhao X, Li J, Niu P, Yang B, Wu H, Wang W, Song H, Huang B, Zhu N, Bi Y, Ma X, Zhan F, Liang Wang, Tao Hu, Hong Zhou, Zhenhong Hu, Weimin Zhou, Li Zhao, Jing Chen, Yao Meng, Ji Wang, Yang Lin, Jianying Yuan et al. Genomic characterisation and epidemiology of 2019 novel coronavirus: implications for virus origins and receptor binding. The Lancet. VOLUME 395, ISSUE 10224, P565–574, 2020. DOI: https://doi.org/10.1016/S0140-6736(20)30251-8.

6. Chan JF, Kok KH, Zhu Z, Chu H, To KK, Yuan S, Yuen KY. Genomic characterization of the 2019 novel human-pathogenic coronavirus isolated from a patient with atypical pneumonia after visiting Wuhan. Emerg Microbes Infect. 2020; 9(1):221–236.

7. Lam TT, Shum, M.H., Zhu, H. et al.. Identifying SARS-CoV-2 related coronaviruses in Malayan pangolins. Nature. 2020. https://doi.org/10.1038/s41586-020-2169-0.

8. Pensaert M, Callebaut P, Vergote J. Isolation of a porcine respiratory, non-enteric coronavirus related to transmissible gastroenteritis. Vet Q. 1986. 8:257–261. doi: 10.1080/01652176.1986.9694050.

9. Li W, Shi Z, Yu M, Ren W, Smith C, Epstein JH, et al. Bats are natural reservoirs of SARS-like coronaviruses. Science. 2005; 310 (5748):676–9. doi:10.1126/science.1118391.

10. Hu B, Ge, X, Wang L et al. Bat origin of human coronaviruses. Virol J 2015; 12, 221. https://doi.org/10.1186/s12985-015-0422-1

11. Graham RL, Baric RS. Recombination, Reservoirs, and the Modular Spike: Mechanisms of Coronavirus Cross-Species Transmission. Journal of Virology Mar. 2010; 84 (7) 3134–3146; DOI: 10.1128/JVI.01394-09.

12. Li F. Structure, Function, and Evolution of Coronavirus Spike Proteins. Annu Rev Virol. 2016; 3(1):237–261. doi:10.1146/annurev-virology-110615-042301

13. Hilgenfeld R, Peiris M. From SARS to MERS: 10 years of research on highly pathogenic human coronaviruses. Antiviral Res. 2013; 100:286–95.

14. Gao H, Yao H, Yang S, Li L. From SARS to MERS: evidence and speculation. Front Med. 2016; 10: 377–82.

15. Coleman CM, Frieman MB. Emergence of the Middle East respiratory syndrome coronavirus. PLoS Pathog. 2013; 9:e1003595.

16. Zhou, P., Yang, X., Wang, X. et al. A pneumonia outbreak associated with a new coronavirus of probable bat origin. Nature. 2020; 579, 270–273. https://doi.org/10.1038/s41586-020-2012-7.

17. Wu, F., Zhao, S., Yu, B. et al. A new coronavirus associated with human respiratory disease in China. Nature. 2020; 579, 265–269. https://doi.org/10.1038/s41586-020-2008-3

18. Wong MC, Cregeen SJJ, Ajami NJ, Petrosino JF. Evidence of recombination in coronaviruses implicating pangolin origins of nCoV-2019. bioRxiv. 2020.

19. Xiao K, Zhai J, Feng Y, Zhou N, Zhang X, Zou J-J, et al. Isolation and Characterization of 2019-nCoV-like Coronavirus from Malayan Pangolins. bioRxiv. 2020. 2020:2020.02.17.951335. doi: 10.1101/2020.02.17.951335.

20. Lam TT-Y, Shum MH-H, Zhu H-C, Tong Y-G, Ni X-B, Liao Y-S, et al. Identification of 2019-nCoV related coronaviruses in Malayan pangolins in southern China. bioRxiv. 2020:2020.02.13.945485. doi: 10.1101/2020.02.13.945485.

21. Zhang YZ, Holmes EC. A Genomic Perspective on the Origin and Emergence of SARS-CoV-2. Cell. 2020; 181(2):223–227. doi:10.1016/j.cell.2020.03.035.

22. Stevens EL, Timme R, Brown EW, et al. The Public Health Impact of a Publically Available, Environmental Database of Microbial Genomes. Front Microbiol. 2017;8:808. Published 2017 May 9. doi:10.3389/fmicb.2017.00808.

23. Sievers F, Wilm A, Dineen D, Gibson J, Karplus K, Li W, Lopez R, McWilliam H, Remmert M, Söding J, Thompson D. and Higgins G. Fast, scalable generation of high-quality protein multiple sequence alignments using Clustal Omega. Mol. Syst. Biol. 2011; 7:539.

24. Rozas J, Ferrer-Mata, A, Sánchez-DelBarrio C, Guirao-Rico S, Librado P, Ramos-Onsins E, Sánchez-Gracia A. DnaSP 6: DNA Sequence Polymorphism Analysis of Large Datasets. Mol. Biol. Evol. 2017; 34: 3299–3302.

25. Bandelt H-J, Forster P, Röhl A. Median-joining networks for inferring intraspecific phylogenies. Mol Biol Evol. 1999; 16:37–48

26. Kühnert D, Stadler T, Vaughan TG, Drummond AJ. Simultaneous reconstruction of evolutionary history and epidemiological dynamics from viral sequences with the birth–death SIR model. 2014. Journal of the Royal Society Interface, 11(94), 20131106.

27. Stadler T, Kühnert D, Bonhoeffer S, Drummond AJ. Birth–death skyline plot reveals temporal changes of epidemic spread in HIV and hepatitis C virus (HCV). 2013. Proceedings of the National Academy of Sciences, 110(1), 228–233.

28. Li R, Pei S, Chen B, Song Y, Zhang T, Yang W, et al. Substantial undocumented infection facilitates the rapid dissemination of novel coronavirus (COVID-19) medRxiv. 2020. https://www.medrxiv.org/content/early/2020/02/17/2020.02.14.20023127.

29. Kucharski AJ, Russell TW, Diamond C et al. Early dynamics of transmission and control COVID-19: a mathematical modelling study. Lancet Infect Dis. 2020; (published online March 11.) https://doi.org/10.1016/S1473-3099(20)30144-4

30. Wang M, Jiang A, Gong L, Luo L, Guo W, Li C, Zheng J, Li C, Yang B, Zeng J, Chen Y, Zheng K, Li H. Temperature significant change COVID-19 Transmission in 429 cities. medRxiv. 2020. doi: https://doi.org/10.1101/2020.02.22.20025791

31. Pawar S, Stanam A, Chaudhari M, Rayudu D. Effects of temperature on COVID-19 transmission. medRxiv. doi: https://doi.org/10.1101/2020.03.29.20044461

32. Prem K, Liu Y, Russell TW, Kucharski AJ, Eggo RM, Davies N. The effect of control strategies to reduce social mixing on outcomes of the COVID-19 epidemic in Wuhan, China: a modelling study. Lancet Public Health. 2020; 5: e261.#x2013;70. 2020. https://doi.org/10.1016/S2468-2667(20)30073-6

